# Playing-related physical problems: a large-scale online survey of professional and amateur Japanese drummers

**DOI:** 10.64898/2025.12.22.25342175

**Authors:** Satoshi Yamaguchi, Kazuaki Honda, Shizuka Sata, Mizuki Komine, Izumi Sakamoto, Makio Kashino, Shinya Fujii

**Affiliations:** Graduate School of Media and Governance, Keio University, 5322 Endo, Fujisawa, Kanagawa, 252-0882, Japan; Communication Science Laboratories, NTT, Inc., 3-1 Morinosato Wakamiya, Atsugi, Kanagawa, 243-0124, Japan; Factor-Inwentash Faculty of Social Work, University of Toronto, 246 Bloor Street West, Toronto, Ontario, M5S 1V4, Canada; Faculty of Environment and Information Studies, Keio University, 5322 Endo, Fujisawa, Kanagawa, 252-0882, Japan; Keio University Research Center for Music Science, Keio University Global Research Institute (KGRI), 2-15-45 Mita, Minato-ku, Tokyo, 108-8345, Japan

## Abstract

Musical performances are generated through bodily movements; because they involve repetitive physical actions, there is a risk of developing playing-related physical problems (PRPPs). This study aimed to investigate PRPPs among amateur and professional drummers in Japan using an online survey, focusing on the differences between the four limbs. In addition, the study sought to compare the prevalence, affected body regions, and diagnoses of PRPPs between the two groups and identify factors associated with the development of PRPPs. The questionnaire included items regarding age, sex, handedness, musical genre, performance level, practice time, experience with PRPPs, affected body regions, and medical diagnoses. Data from 868 respondents (667 amateurs and 201 professionals) were analyzed. The self-reported prevalence of PRPPs was 33.4% and 66.2% in amateurs and professional, respectively. Among amateurs, the symptoms were most frequent in the right wrist, whereas among professionals, the right lower limb was most frequently affected. Regarding diagnosis, the most frequently reported condition was tenosynovitis among amateurs (2.10 %) and musician’s dystonia among professionals (8.95%). Logistic regression analysis revealed that the risk factors for PRPPs included higher performance levels, more significant stress related to the use of the metronome click, perfectionism, and experience of altering playing technique. In contrast, increased stress resilience was significantly associated with a decreased risk of PRPP development. There was a significant interaction indicating that the protective effect of stress resilience weakened at higher performance levels. This study revealed that professional drummers exhibited a higher prevalence of PRPPs than did amateurs, with distinct patterns of affected regions and diagnoses. Additionally, the performance level, psychological factors, and playing environment may contribute to these problems. This study may provide evidence for the revision of practice and performance methods for injury prevention tailored to each performance level.

## Introduction

Musical performance requires repetitive body movements, with the risk of playing-related physical problems (PRPPs) [1–3]. Playing-related musculoskeletal disorders (PRMDs) and musician’s dystonia (MD) have been identified as significant issues associated with PRPPs [3]. Zaza et al. defined PRMDs as “any pain, weakness, numbness, tingling, or other symptoms that interfere with your ability to play your instrument at the level you are accustomed to” [4]. Meanwhile, MD is a type of task-specific dystonia characterized by involuntary muscle contractions occurring during instrumental performance [5]. PRPPs can significantly impair skilled motor control in musicians and may become a major factor hindering the long-term continuation of their musical careers [6,7]. In recent years, a number of Japanese drum kit players (drummers) from well-known pop/rock bands have reported the development of MD and associated issues, rest periods, and retirements. Considering that professional musicians often begin their careers as amateurs, investigating PRPPs among amateur players is essential for developing preventive educational strategies and sustaining musicians’ careers.

Previous questionnaire surveys of university music students and orchestra musicians indicated that prolonged practice and performance under highly stressful conditions are significant risk factors for PRPPs and that the affected body regions vary depending on the instrument [6–10]. For instance, in MD, the body locations affected are often related to technical demands, such as the right hand for keyboard and plucked string players and the left hand for string players [11]. Sandell et al. conducted a large-scale survey of PRMDs among percussionists (n = 279), who were primarily classical players [12]; the most affected body locations were the left and right hands (46.4% and 44.6%, respectively) and the left and right hips (38.7% and 39.3%, respectively), indicating that many participants experienced multiple PRMDs. In addition, Sandell et al. highlighted the potential role of stress in increasing the risk of PRMDs, given that 76.9% of the affected musicians reported high levels of stress.

Drumming is no exception, and it is a physically demanding activity [13,14]. Because of the seated playing posture, simultaneous use of all four limbs, repetitive movements, and need for reaching, drummers often adopt non-neutral body postures. Regarding musical genre, unlike classical percussionists, many drummers play other musical genres (e.g., rock, pop, and jazz), which are more improvisational than classical music. However, nowadays, even drummers playing rock and pop music experience musical constraints that differ from those in the past due to stage performance evolution [15]. Nowadays, in rock and pop music, live performances often require synchronization with videos, stage lighting, special effects, and prerecorded tracks. This places a high demand on drummers to play accurately while listening to the metronome click and maintain the overall ensemble of the band. Professional drummers are required to maintain strict temporal accuracy, even in large stages, where the slightest deviation is unacceptable. This demands sustained concentration and puts drummers under high pressure, leading to the accumulation of mental strain and stress.

Notably, Altenmüller and Jabusch identified several demanding musical constraints as potential factors contributing to a higher incidence of MD among classical musicians than among musicians in other genres [7]. These include the requirement for temporal precision at the millisecond level, intense pressure arising from audience and peer expectations, a performance environment that allows for no margin of error, and elevated levels of performance anxiety and stress. In recent years, all famous Japanese drummers with a reported onset of MD and the accompanying struggles, breaks, and retirement have been members of bands capable of performing solo concerts in arenas that can accommodate more than 10,000 people, suggesting that they may be in a situation similar to that of classical musicians. Drummers who perform under extremely high musical constraints have reported MD, and such circumstances among professional drummers may be associated with the onset of PRPPs. However, related studies have mainly focused on percussionists with classical music training, and research on PRPPs in drummers remains extremely limited [13,14,16–18].

To address this knowledge gap, Azar conducted a large-scale online questionnaire survey of drummers and a detailed analysis of the prevalence and risk factors of PRMDs among drummers [13]. The survey was conducted using snowball sampling via social media, and valid responses were collected from 831 drummers across 49 countries. The results showed a self-reported lifetime PRMD prevalence rate of 68%; among the patients, 59% were had experienced multiple disorders. The most commonly affected region was the upper limbs (59%), with particularly high incidence of wrist problems (25%). Tendinitis, tenosynovitis, carpal tunnel syndrome, and arthritis were the most common diagnoses reported, suggesting that the large force applied during drumming, vibration caused by the impact of hitting, highly repetitive nature of the work, long duration of the same posture, and non-natural posture may be factors affecting the onset of PRMDs in drummers. Furthermore, of the participants who had experienced PRMDs, only 42% had received a diagnosis at a medical institution, highlighting the fact that many drummers did not receive a formal diagnosis or appropriate treatment.

Based on Azar’s study [13], however, further detailed analyses are needed from three perspectives. First, data from Japan are lacking. In Azar’s study, 87.8% of participants were from North America and Europe, whereas only 8.8% were from the Asia-Pacific region, including Japan. Recent research on music has focused on Western musicians, and this overrepresentation of Western, Educated, Industrialized, Rich, and Democratic (“WEIRD”) societies has been identified as a factor that hinders analyses of global cultural diversity [19]. According to the *Global Music Report 2025* published by the International Federation of the Phonographic Industry (IFPI), Japan’s music market was the second largest in the world in 2024, following the United States, a position it has maintained for over a decade [20,21]. Therefore, professional drummers in Japan are likely to have more opportunities to perform in synchrony with staging in large-scale live venues compared to drummers in other countries. Given that performance environments characterized by high musical constraints may increase the psychological burden and stress of drummers and potentially contribute to the development of PRPPs, it is imperative to collect data from drummers in Japan.

Second, professional and amateur drummers should be compared. In Azar’s study [13], participants were limited to those who played drums for more than 5 h per week, resulting in a sample composed primarily of experienced professional drummers. The study reported that participants had an average of 26 ± 15 years of playing experience, and 80% of them performed as professionals. Amateur drummers who played drums recreationally and did not meet this criterion were excluded from the study, making it difficult to comprehensively assess the risk and prevalence rates of PRPPs among amateur and professional drummers. Beveridge et al. [22] studied tapping movements in drummers at different levels and reported that amateur drummers exhibited higher co-contraction between the wrist flexors and extensors, indicating insufficient coordinated control of antagonistic muscle groups. The results of the study suggest that higher co-contraction leads to increased muscle loading in amateur drummers than in professional drummers, and amateur drummers can be at risk of developing PRMDs, such as muscle fatigue and motor dysfunction. In contrast, although professional drummers may have acquired efficient performance techniques that minimize unnecessary muscle loading through years of training, they face different types of risks, namely, psychological pressure associated with the expectation of flawless performance in high-stakes situations, such as playing in large concert venues before a vast audience or recording in commercial settings. MD prevalence is higher among professionals than in amateurs, possibly because of the cumulative effects of long-term performance experience and psychological pressure to deliver a flawless performance [9]. This finding suggests that the risk of PRPPs associated with MD is higher among professional musicians.

Third, there is a lack of a detailed analysis of the differences among the four limbs. In drum kit performance, the four limbs are required to move independently, resulting in different roles for each limb. Generally, a right-handed drummer plays a hi-hat with their right hand, a snare drum with their left hand, a bass drum with their right foot, and a hi-hat pedal with their left foot. For example, playing a hi-hat with the right hand often involves fine subdivisions that require motor control at high frequencies. In contrast, although the left hand playing the snare drum and the right foot playing the bass drum are less frequently used for fine subdivisions than the right hand, the left hand and right foot play a central role in maintaining rhythmic patterns, requiring coordination between sensory and motor processes and accurate timing control. Owing to these differences in movement, the technical demands placed on each limb vary. As mentioned previously, MD tends to develop in body parts with higher technical demands. Furthermore, Jabusch and Altenmüller reported that it tends to affect the dominant hand [23]. Considering these factors, right-handed drummers may be at a higher risk of developing MD in their right hand. Furthermore, given that temporal sensorimotor constraints, such as imbalances in the use of the hands, affect the onset of MD [7], there may be a risk of onset in the left hand (snare drum) or right foot (bass drum). Based on these biomechanical characteristics of drum performance, a detailed analysis of the differences between the four limbs is expected to deepen our understanding of PRPPs in drummers.

Therefore, this study aimed to directly compare PRPPs between amateur and professional drummers in Japan. We examined the self-reported prevalence of PRPPs, affected body regions, and medical diagnoses by comparing these variables between amateur and professional groups to investigate the factors contributing to the development of PRPPs.

## Materials and Methods

### Research design and participants

This study targeted drummers aged ≥18 years residing in Japan, regardless of musical genre. Eligible participants included individuals who played drums as a personal hobby, individuals taking private lessons, members of amateur bands, students enrolled in music or technical colleges, part-time or full-time drum instructors, part-time or full-time session drummers, and drummers associated with one or more bands. Musicians who primarily played other instruments were included if they had experience playing drums. Drummers who were not actively playing at the time of recruitment were included, provided they had prior drumming experience. The criteria were intended to include individuals who had been forced to retire from playing due to PRPPs. We defined amateur and professional drummers based on the following question [24]:

“Please indicate the music-activity level that best applies to you, either present or past.”

- Pro (For pay, national level)
- Pro (For pay, local level)
- Amateur (No pay, have performed in paid shows)
- Amateur (No pay, have performed in free shows)
- Amateur (No pay, enjoy playing on my own)
- No experience

All procedures were approved by the Research Ethics Committee of Keio University (No. 376) and the Research Ethics Committee of NTT Communication Science Laboratories (R03-013). Informed consent was obtained from all the participants. This study was conducted and reported in accordance with the Strengthening the Reporting of Observational Studies in Epidemiology (STROBE) guidelines for cross-sectional studies (S1 File).

### Survey items

We developed an online questionnaire based on a thorough review of the literature [7,11,15,20,25,26] and consultations with three professional drummers who had experienced PRPPs (S2 File: the original questionnaire in Japanese; S3 File: the questionnaire translated in English). To capture overall trends from a larger number of participants, we designed a simplified questionnaire items using a 5-point Likert scale instead of existing assessment scales. The questionnaire included the following items: age; sex; handedness; ratio of matched grip to traditional grip usage; genre of music; favorite musician; performance level (amateur or professional); degree of perfectionism; degree of overfocusing; degree of stress resilience; sleep quality; degree of musical improvisation and constraints in performed music; starting age of drumming and total years of experience; total hours of individual practice; experience and starting age of formal lessons; total hours of formal music lessons; content of individual practice; experience of change of play style; starting age and total years of playing another instrument; experience with and starting age of sports activities; usage of the click tracks; degree of stress related to the click tracks; presence and details of PRPPs (including related episodes); affected body regions; and experience of medical diagnosis and names of diagnosis.

### Original items on potential risk factors

This section examines how risk factors for PRPPs previously identified in studies on musicians relate to PRPPs among drummers. To capture the overall trends from a larger number of participants, we used a simplified questionnaire items with a 5-point Likert scale rather than existing assessment scales.

- I am a perfectionist (I am not tolerant to even the slightest mistakes, tend to focus on flaws or past failures rather than successes, etc.) [7,27]
- I tend to overfocus even on the small details (overfocusing) [27,28]
- I am resilient to stress (stress resilience) [25,29]
- I get enough sleep [26]
- The genre of music I play is open to improvisation. [7,30]
- I often play to sound sources other than live performances (e.g., click tracks, soundtracks, programmed music, etc.) [7,30]

Participants who reported experiencing PRPPs could select multiple affected body locations from a body map divided into 55 detailed areas (S1 Fig). Furthermore, participants who experienced multiple types of PRPPs were allowed to separately report each instance by repeating the same procedure. Five drummers piloted the questionnaire to ensure that it functioned in an online environment and to obtain feedback on its ease of use, clarity, and burden on respondents. The questionnaire was designed to collect data anonymously.

### Participant recruitment

The participants were recruited using snowball sampling on social media and via a magazine. The authors posted URL for the questionnaire on their social media accounts (Facebook, Instagram, and X [formerly Twitter]). SY and SF published an article requesting for study participation, both in print and online, through an external outlet, *Rhythm & Drums Magazine*, which is the most widely used drum-focused media source in Japan. The article primarily mentioned the experience of SY and included a URL to the questionnaire. Additionally, we made recruitment efforts through communities of drummers’ and musicians,’ including universities and private music schools. Based on a previous study [9], the target sample size was set at 1,000 respondents. The survey period was set from March 2, 2022, to June 30, 2022. To ensure data reliability, we set a trap question and excluded respondents who checked it.

### Procedures

Participants who accessed the questionnaire website were first required to review the study information sheet; only those who provided consent were allowed to complete the questionnaire. They were informed that they could withdraw from the survey at any time by closing their browser during the response process. In addition, we ensured that even after completing the questionnaire, their responses could be deleted upon request by contacting the researchers later. Participant confidentiality was maintained by assigning a research-specific ID that did not allow for personal identification, and all data were handled in an anonymized form.

### Data analysis

Demographic and PRPP data were analyzed using descriptive statistics for continuous variables and response frequencies for categorical variables. Independent *t*-tests were used to compare continuous variables, and chi-square tests were used to compare categorical variables. Based on the diagnostic classification method used in Azar’s study [13], a physical therapist (SS) conducted a qualitative analysis in which several specific diagnostic terms were grouped under a single diagnostic category.

Logistic regression analysis was conducted to identify PRPP-related factors. Respondents with missing values were excluded. Numerical variables were standardized to a mean of 0 and SD of 1. A stepwise selection method based on the Akaike information criterion (AIC) was used to determine the model for analysis. An initial null model containing only the intercept was constructed; a full model was set up to examine the main effects of the variables considered relevant and the interaction terms with Level to assess whether the effects of these variables differed across music-activity levels.

After estimating the coefficients, odds ratios for each variable were calculated to evaluate the influence of each effect. The variance inflation factor (VIF) was used to check for multicollinearity. We evaluated the predictive performance of the model by calculating the area under the receiver operating characteristic (ROC) curve (AUC) and Nagelkerke’s R^2^ using the R packages (“pROC” [31] and “performance” [32]). AUC value was presented with a bootstrap 95% confidence interval with bootstrapping techniques based on 2,000 bootstrapped samples. In addition, we performed a 10-fold cross-validation procedure to examine the degree of overfitting in the selected models. All statistical analyses were performed using the R software (ver. 4.4.1), with the significance level of *p* < 0.05.

For participants who reported stress related to the use of a metronome click, underlying causes were explored through an open-ended survey question: *“Have you ever felt stress by using a click track? If you have, describe the situation where you felt stressed in the space below (e.g., I felt stressed when only I had to listen to a click track during a live performance. It required a lot of concentration, because being out of sync with the click track means that I am not in sync with the lighting, video, and other production elements).”* The responses to this question were analyzed using reflexive thematic analysis [33] following an inductive, data-driven approach. Two researchers (SY and IS) independently conducted the initial coding using NVivo (QSR International), a qualitative data analysis software. Intercoder dialogue was used to compare interpretations, discuss discrepancies, and refine the coding framework to ensure analytical depth and credibility. The final codes were exported to Microsoft Excel for further organization and visualization of thematic patterns. The initial analysis was conducted in Japanese language to retain the original nuances of the responses. After synthesis and organization of the themes, they were translated into English.

The researcher’s positionality informed the analytical process. The lead researcher (SY) is a professional drummer with lived experience of MD and has personally encountered stress related to metronome click use in professional performance contexts. The other researcher (IS) who analyzed the qualitative data has over 20 years of experience in qualitative research and is an amateur drummer. These distinct yet complementary perspectives enriched the interpretive process and supported a reflexive engagement with the data.

## Results

### Demographics of the respondents

Ultimately, 1,099 responses were obtained. After excluding respondents aged <18 years, those who failed to respond appropriately to the trap question, and those with incomplete responses (n = 231), data from 868 participants (667 amateurs and 201 professionals) were included in the analysis (Table 1). Self-identified sex was reported by 688 males (78.57%), 172 females (19.82%), and 14 non-binary individuals or non-respondents (1.61%). The mean age of all the participants was 29.58 years (standard deviation [SD]: 10.89, range: 18–65 years), as shown in Table 2. Among amateurs, the mean age was 27.55 years (SD: 10.11, range: 18–65), whereas it was 36.34 years (SD: 10.69, range: 18–65) among professionals. Regarding handedness, 788 participants were right-handed (90.78%), 55 participants were left-handed (6.34%), and 25 participants were ambidextrous (2.88%).

**Table 1.**
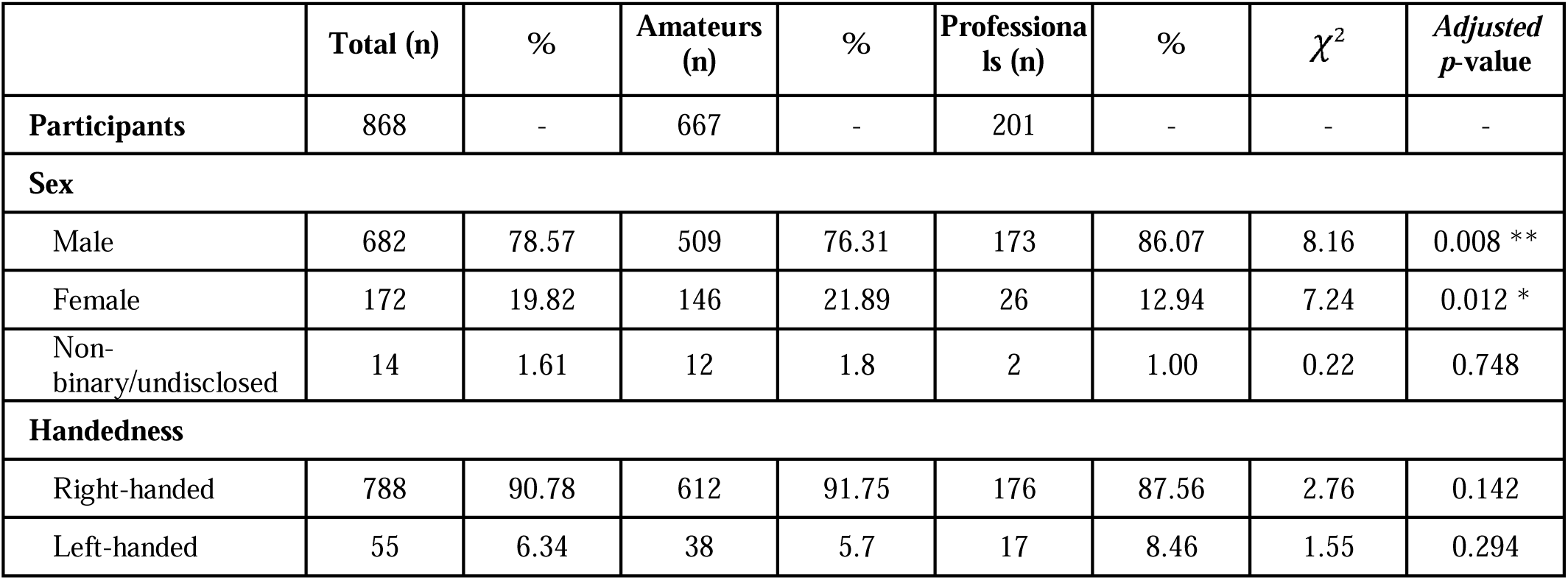
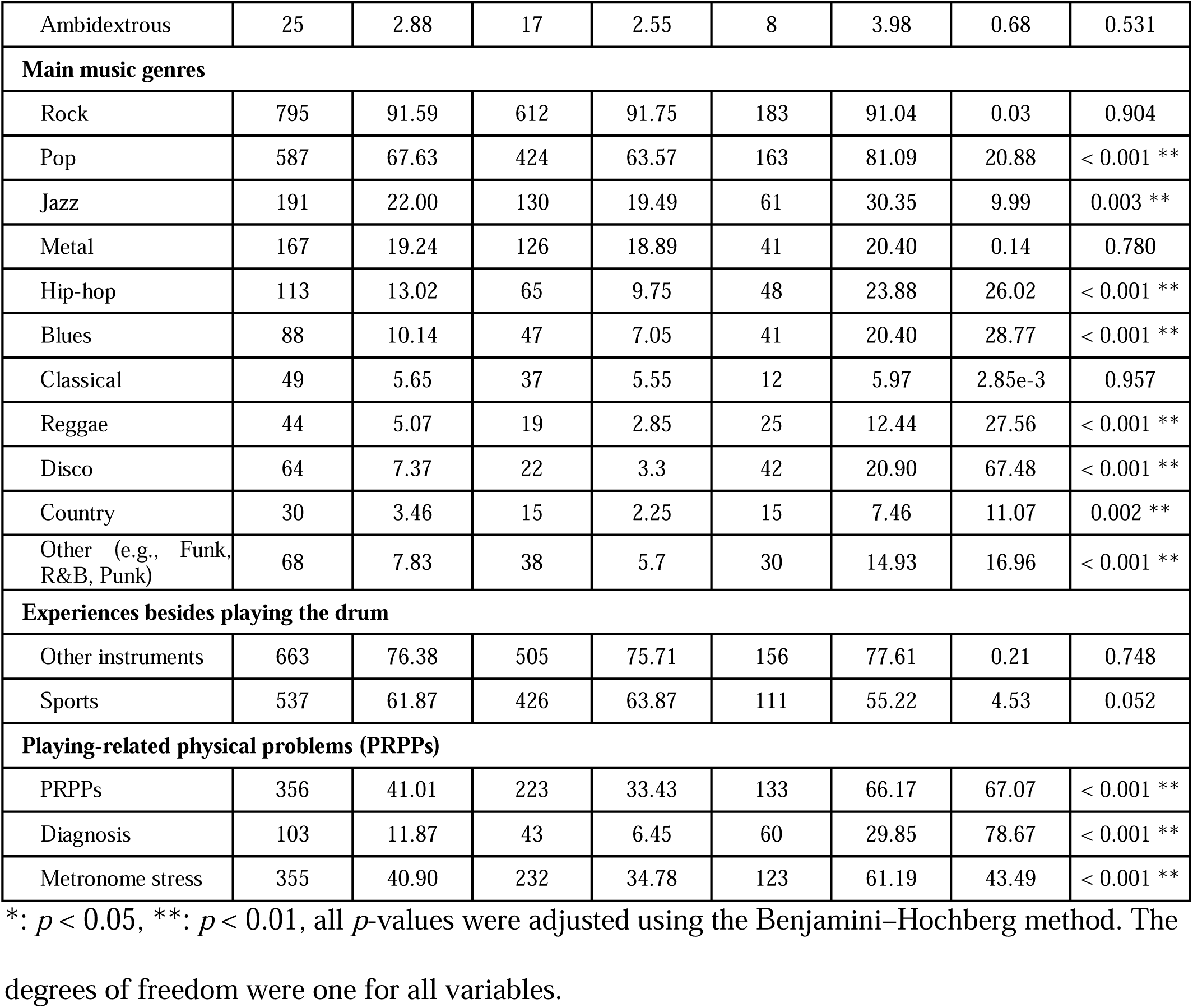
Participant Demographics: Categorical Variables.

| | Total (n) | % | Amateurs (n) | % | Professionals (n) | % | $\chi^2$ | Adjusted *p*-value |
| --- | --- | --- | --- | --- | --- | --- | --- | --- |
| **Participants** | 868 | - | 667 | - | 201 | - | - | - |
| **Sex** |  |  |  |  |  |  |  |  |
| Male | 682 | 78.57 | 509 | 76.31 | 173 | 86.07 | 8.16 | 0.008 ** |
| Female | 172 | 19.82 | 146 | 21.89 | 26 | 12.94 | 7.24 | 0.012 * |
| Non-binary/undisclosed | 14 | 1.61 | 12 | 1.8 | 2 | 1.00 | 0.22 | 0.748 |
| **Handedness** |  |  |  |  |  |  |  |  |
| Right-handed | 788 | 90.78 | 612 | 91.75 | 176 | 87.56 | 2.76 | 0.142 |
| Left-handed | 55 | 6.34 | 38 | 5.7 | 17 | 8.46 | 1.55 | 0.294 |
| Ambidextrous | 25 | 2.88 | 17 | 2.55 | 8 | 3.98 | 0.68 | 0.531 |
| **Main music genres** |  |  |  |  |  |  |  |  |
| Rock | 795 | 91.59 | 612 | 91.75 | 183 | 91.04 | 0.03 | 0.904 |
| Pop | 587 | 67.63 | 424 | 63.57 | 163 | 81.09 | 20.88 | < 0.001 ** |
| Jazz | 191 | 22.00 | 130 | 19.49 | 61 | 30.35 | 9.99 | 0.003 ** |
| Metal | 167 | 19.24 | 126 | 18.89 | 41 | 20.40 | 0.14 | 0.780 |
| Hip-hop | 113 | 13.02 | 65 | 9.75 | 48 | 23.88 | 26.02 | < 0.001 ** |
| Blues | 88 | 10.14 | 47 | 7.05 | 41 | 20.40 | 28.77 | < 0.001 ** |
| Classical | 49 | 5.65 | 37 | 5.55 | 12 | 5.97 | 2.85e-3 | 0.957 |
| Reggae | 44 | 5.07 | 19 | 2.85 | 25 | 12.44 | 27.56 | < 0.001 ** |
| Disco | 64 | 7.37 | 22 | 3.3 | 42 | 20.90 | 67.48 | < 0.001 ** |
| Country | 30 | 3.46 | 15 | 2.25 | 15 | 7.46 | 11.07 | 0.002 ** |
| Other (e.g., Funk, R&B, Punk) | 68 | 7.83 | 38 | 5.7 | 30 | 14.93 | 16.96 | < 0.001 ** |
| **Experiences besides playing the drum** |  |  |  |  |  |  |  |  |
| Other instruments | 663 | 76.38 | 505 | 75.71 | 156 | 77.61 | 0.21 | 0.748 |
| Sports | 537 | 61.87 | 426 | 63.87 | 111 | 55.22 | 4.53 | 0.052 |
| **Playing-related physical problems (PRPPs)** |  |  |  |  |  |  |  |  |
| PRPPs | 356 | 41.01 | 223 | 33.43 | 133 | 66.17 | 67.07 | < 0.001 ** |
| Diagnosis | 103 | 11.87 | 43 | 6.45 | 60 | 29.85 | 78.67 | < 0.001 ** |
| Metronome stress | 355 | 40.90 | 232 | 34.78 | 123 | 61.19 | 43.49 | < 0.001 ** |

\*: $p < 0.05$ , \*\*: $p < 0.01$ , all $p$ -values were adjusted using the Benjamini–Hochberg method. The degrees of freedom were one for all variables.

**Table 2.**
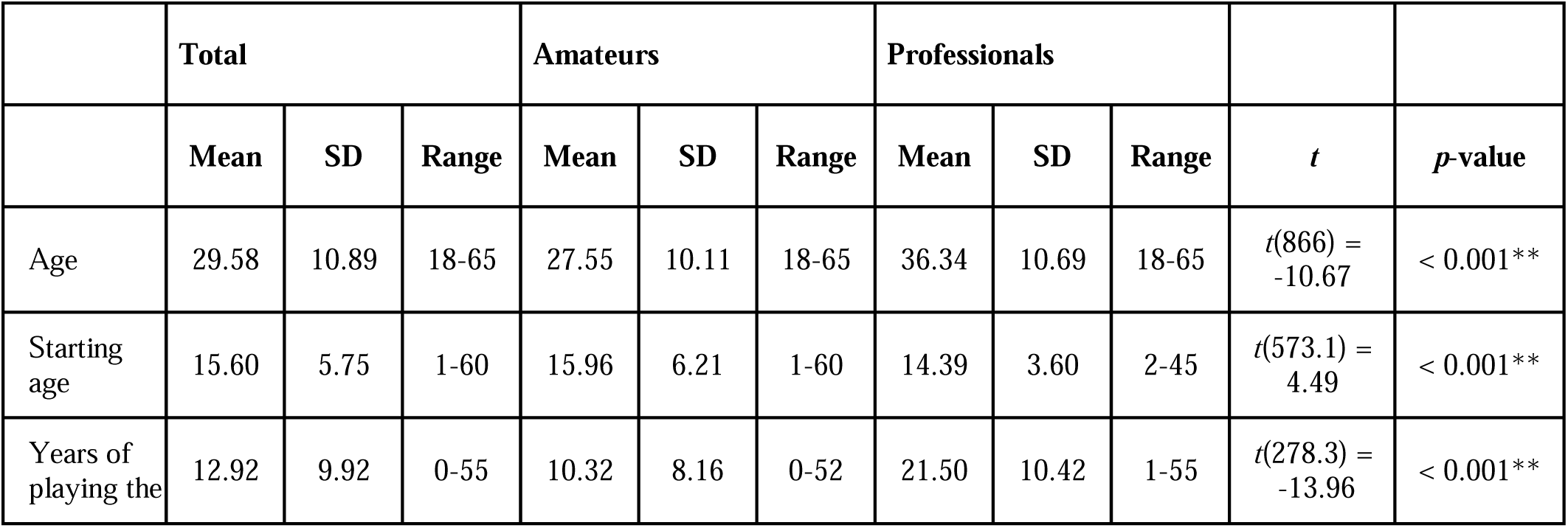
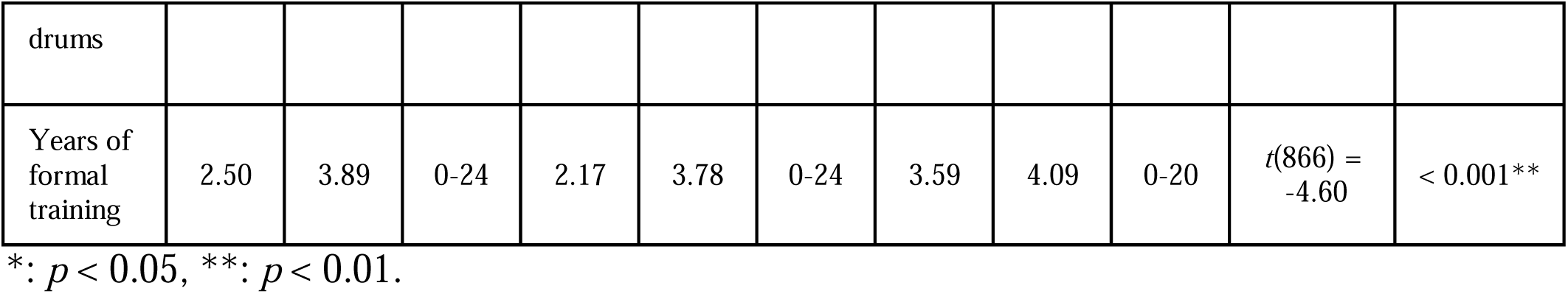
Participant Demographics: Continuous Variables.

The average starting age for drumming was 15.60 years overall (SD: 5.75, range: 1–60 years), as shown in Table 2. Among amateurs, the average was 15.96 years (SD: 6.22, range: 1–60); among professionals, it was 14.39 years (SD: 3.60, range: 2–45). The average number of years of drumming experience was 12.92 years overall (SD: 9.92, range: 1–60). Among amateurs, the average was 10.32 years (SD: 8.16, range: 1–60); among professionals, it was 21.50 years (SD: 10.42, range: 2–45). As one of the musical genres of the respondents, “Rock” was the most frequently reported (n = 795, 91.59%). Most respondents reported their experience with other musical instruments (n = 663, 76.38%) and sports (n = 537, 61.87%). Furthermore, 356 respondents (41.01%) reported their experience with PRPPs, including 223 amateurs (33.43%) and 133 professionals (66.17%). In the PRPPs group, 103 respondents (28.93% of participants with PRPPs) reported having received a medical diagnosis. Stress related to metronome click use was reported by 355 participants (40.90%), including 232 amateurs (34.78%) and 123 professionals (61.19%). Chi-square tests revealed that professional drummers were significantly more likely to report experiencing PRPPs (χ*^²^*(1) = 67.07, *p* < 0.001), having a history of medical consultation for PRPPs (χ*^²^*(1) = 78.67, *p* < 0.001), and experiencing stress related to metronome click use (χ*^²^*(1) = 43.49, *p* < 0.001) than did amateur drummers.

### Affected body regions and diagnosis

Among the respondents with self-reported PRPPs, 206 amateurs and 104 professionals provided information about the affected body regions. Among amateurs, the most frequently reported site was the right wrist (27.18% of those who responded to the body region question; Fig 1), followed by the left wrist (25.24%) and the right hand (20.87%). Among professional drummers, the most frequently reported affected locations were the right ankle (37.90%), right shank (32.26%), and right foot (30.65%). Regarding diagnosis, the reported conditions included: MD (professional: n = 18, 8.95% of the participated professional drummers, amateur: n = 5, 0.75% of the participated amateur drummers, Fig 2), tenosynovitis (n = 20), muscle-related issues such as spasms, tension, tears, or fatigue (n = 13), intervertebral disc problems (n = 11), joint disorders or inflammation (n = 5), tendinitis or tendinosis (n = 3), spinal alignment abnormalities or subluxations (including scoliosis) (n = 3), sciatica or spinal nerve compression (n = 1), other PRMDs (n = 10), other conditions (n = 12), and no diagnosis (n = 18).

**Fig 1.**
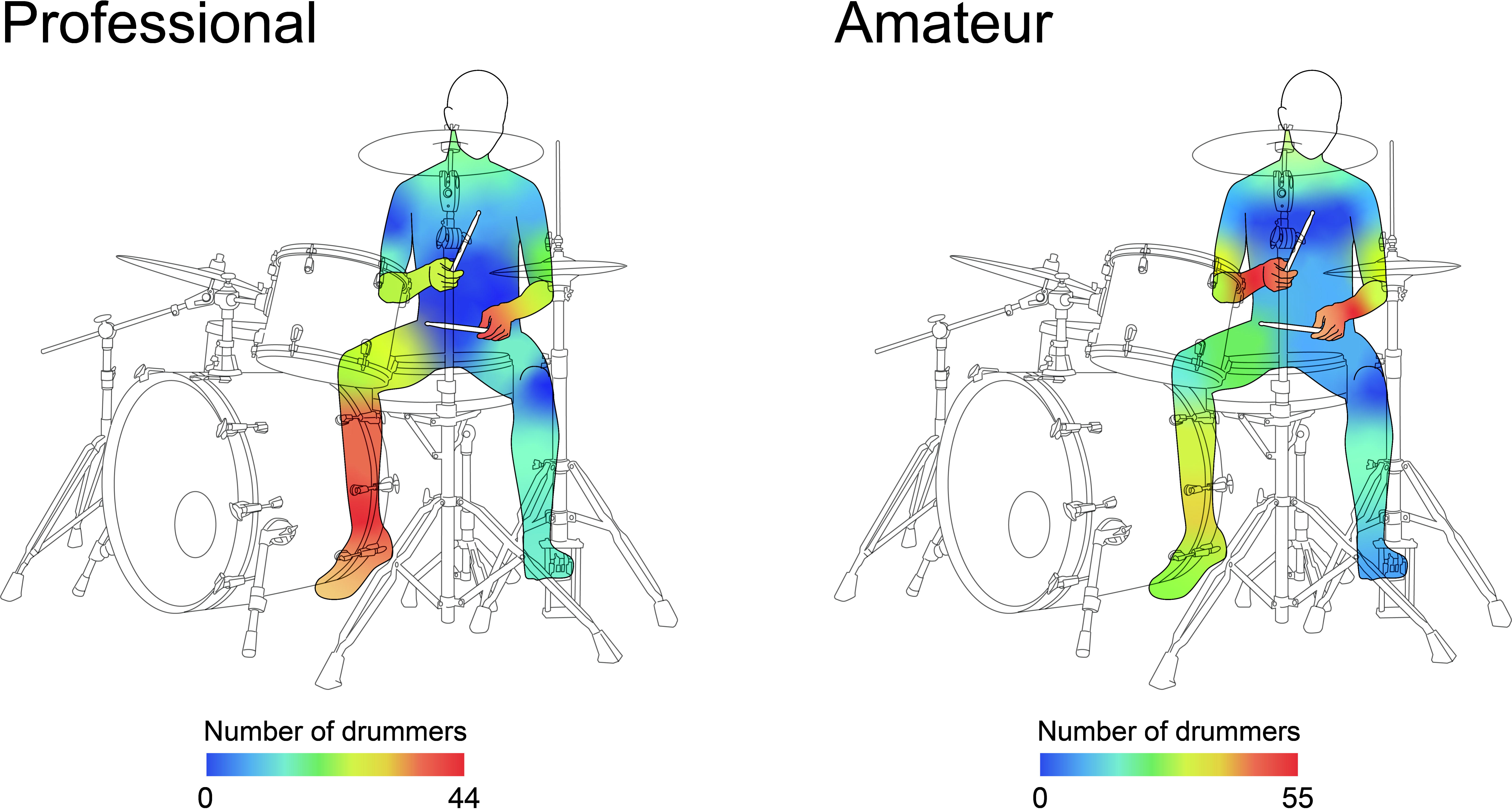
Frequency of affected body regions.

**Fig 2.**
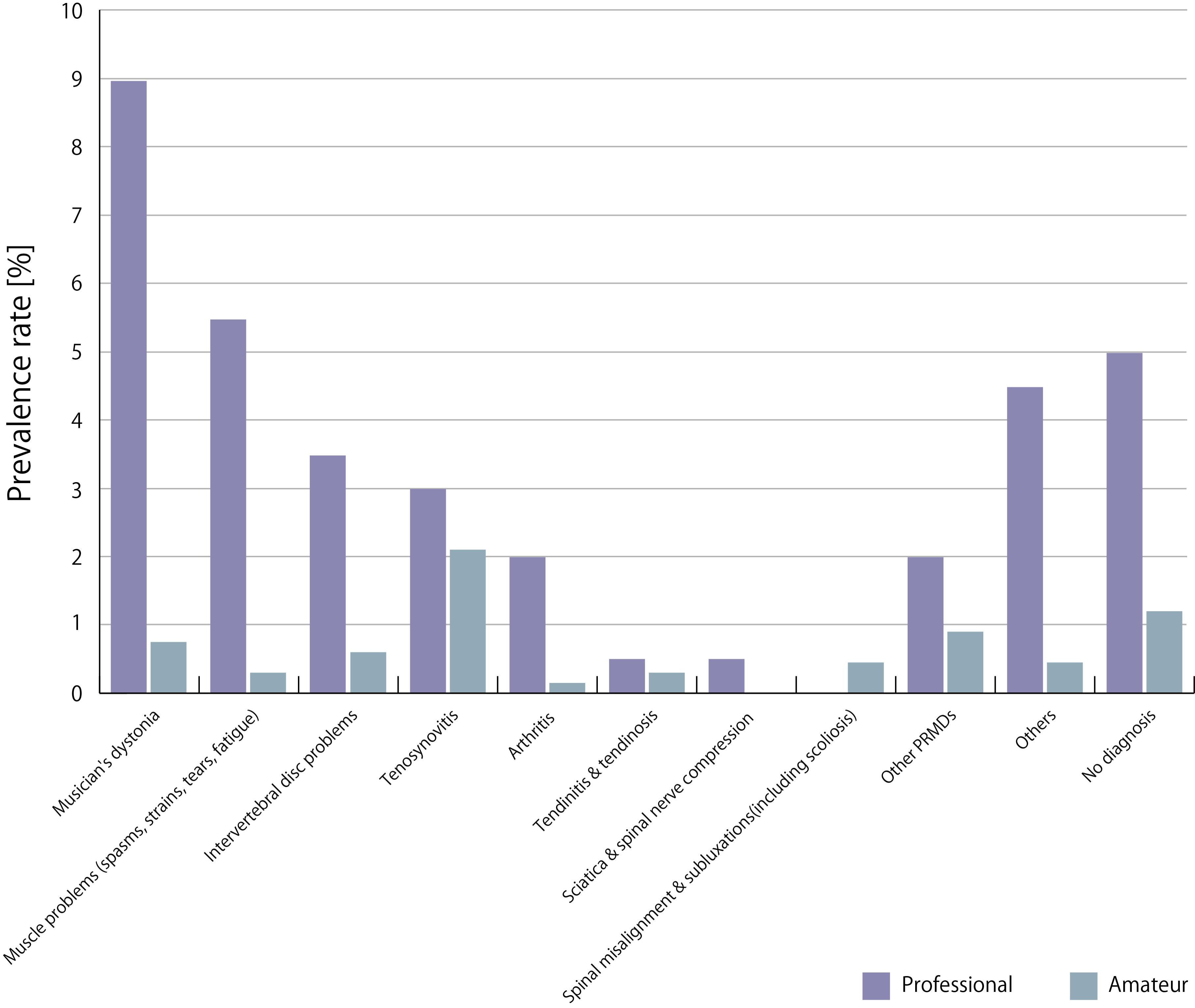
Diagnoses reported among amateur and professional drummers.

Red indicates areas where symptoms were frequently reported, whereas blue indicates areas with fewer reports of symptoms. Based on these responses, the illustrator created an illustration. A heat map generated from the original dataset is shown in S1 Fig.

### Risk factors of MRPPs from logistic regression

The analyzed sample size was reduced from 868 to 813 because of missing values. As a result of model selection, the final model included the main effects of level, metronome stress, perfectionism, stress resilience, change of technique, starting age, experience with Blues music, and experience with sports, as well as the interaction between level and stress resilience (AIC = 992.7). Estimation of the coefficients indicated that increases in level, metronome stress, perfectionism, and change of technique were significantly associated with developing PRPPs, whereas greater stress resilience was associated with a lower probability of developing PRPPs (Table 3). The interaction between level and stress resilience was significant, suggesting that as the level increased, the reducing effect of stress resilience was attenuated. Post hoc analysis revealed that the significant effect of stress resilience disappeared at Levels 3 and 5 (Fig 3).

**Fig 3.**
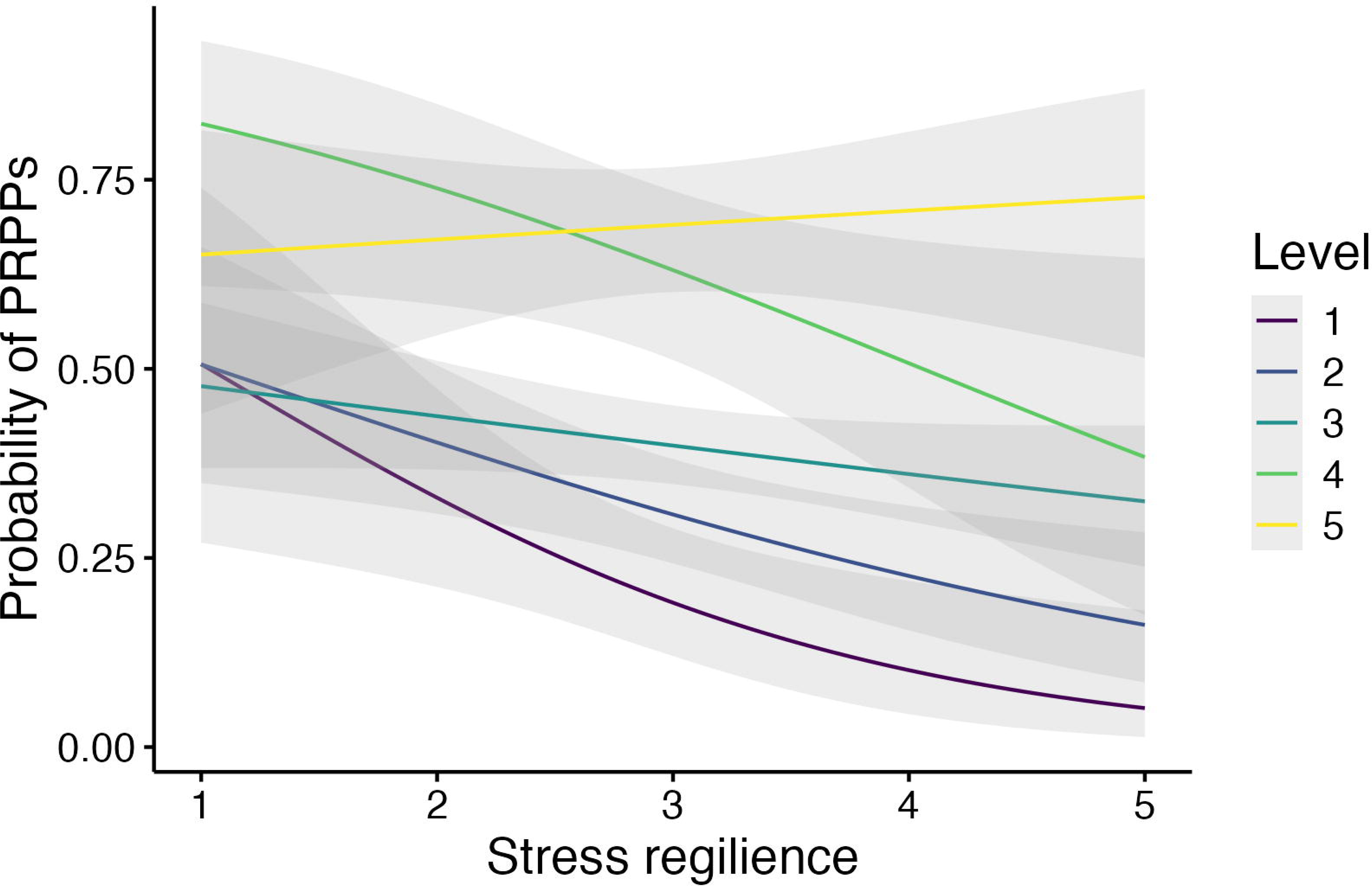
Relationship between PRPPs and stress resilience by level.

**Table 3.** Playing-related physical problems risk factor coefficients from logistic regression.

|  | Estimated coefficient | Standard error | Z-value | p-value | Odds ratio (95% CI) |
| --- | --- | --- | --- | --- | --- |
| (Intercept) | -0.85 | 0.15 | -5.56 | < 0.001 ** | 0.43 (0.31-0.57) |
| Level | 0.62 | 0.09 | 6.87 | < 0.001 ** | 1.86 (1.56-2.22) |
| Metronome stress | 0.64 | 0.16 | 4.02 | < 0.001 ** | 1.90 (1.39-2.61) |
| Perfectionism | 0.26 | 0.08 | 3.25 | 0.001 ** | 1.30 (1.11-1.53) |
| Stress resilience | -0.20 | 0.08 | -2.51 | 0.012 * | 0.82 (0.70-0.96) |
| Change of technique | 0.37 | 0.18 | 2.04 | 0.042 * | 1.45 (1.01-2.08) |
| Starting age | 0.13 | 0.08 | 1.68 | 0.092 | 1.14 (0.98-1.33) |
| Blues | -0.42 | 0.26 | -1.60 | 0.109 | 0.65 (0.39-1.09) |
| Sports | 0.23 | 0.16 | 1.42 | 0.156 | 1.26 (0.92-1.72) |
| Level x Stress resilience | 0.20 | 0.09 | 2.21 | 0.027 * | 1.22 (1.02-1.46) |

\*: $p < 0.05$ , \*\*: $p < 0.01$ .

Levels 1, 2, and 3 indicate amateur drummers, and levels 4 and 5 indicate professional drummers.

A Kruskal–Wallis rank sum test was conducted as a post-hoc analysis, and there was no significant difference in the mean values of stress resilience across levels (χ*²*(4) = 2.31, *p* = 0.680). Therefore, it is unlikely that a ceiling effect occurred in stress resilience at these two levels. The AUC in the final model was 0.73 (*95%CI*: 0.69-0.76), and Nagelkerke R^2^ was 0.20. The result of 10-fold cross-validation showed an AUC of 0.71. All VIFs were <10, and the mean VIF of the model was 1.1.

### Qualitative analysis of self-reported stress associated with metronome usage

In addition to quantitative responses, 223 (34.8%) amateur drummers and 123 (61.2%) professional drummers further responded to an open-ended question on the stress associated with their metronome use (Fig 4). Amateurs frequently reported stress directly associated with the metronome itself, such as difficulty in maintaining synchrony with clicks. In contrast, professionals more often described stress arising from the need to balance the metronome with other concurrent musical demands.

**Fig 4.**
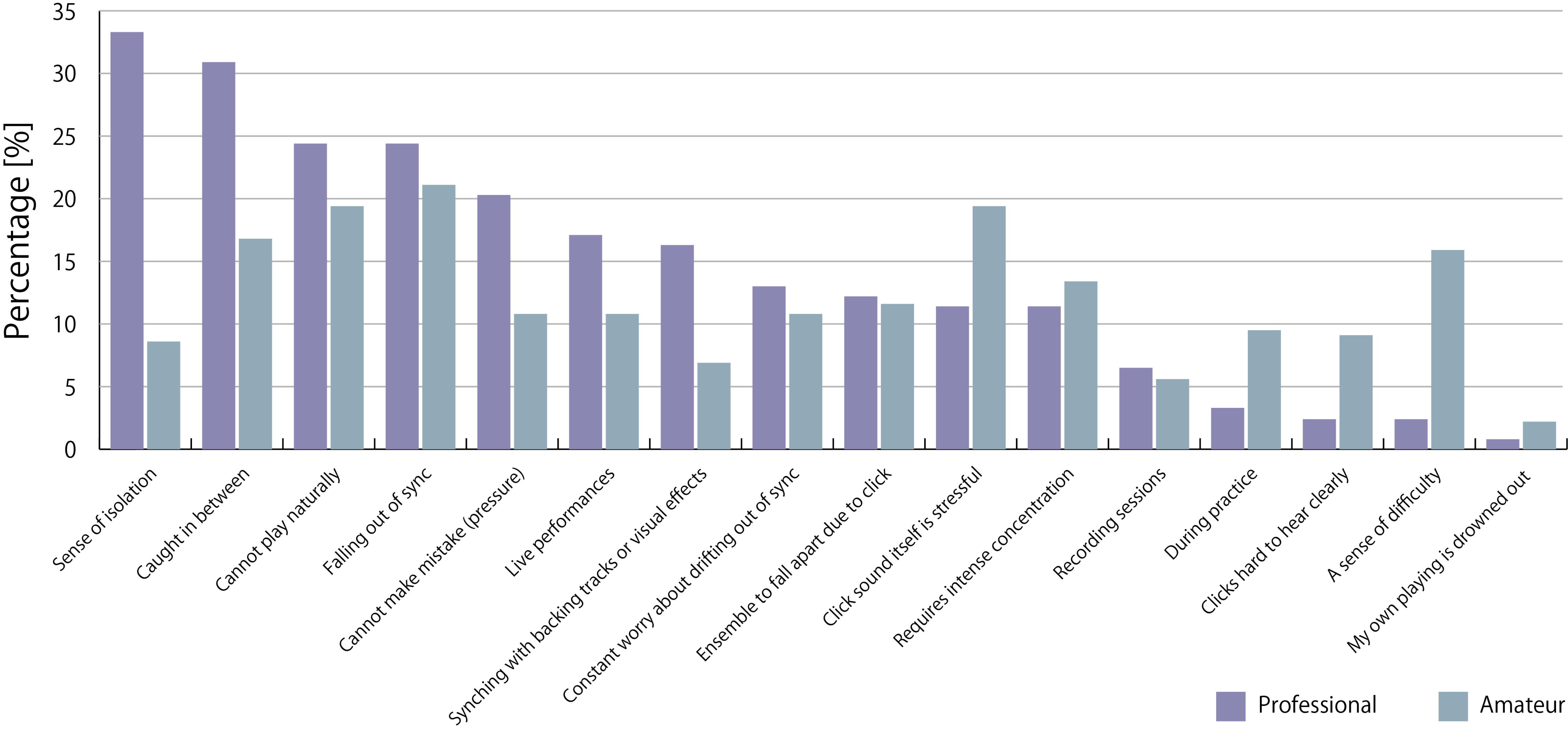
Qualitative analysis of stress experiences during metronome use: a comparison between amateur and professional drummers.

Sense of isolation: sense of isolation (being the only one hearing the click during performance); Caught in between: feeling caught in between the band’s natural groove and click track; Cannot play naturally: unable to play naturally or expressively because of the click track; Falling out of sync: stress when falling out of sync with the click; Cannot make mistake (pressure): pressure and tension from not being allowed to make mistakes; Live performances: stress experienced during live performances; Synching with backing tracks or visual effects: stress from having to sync precisely with backing tracks or visual effects; Constant worry about drifting out of sync: constant worry about drifting out of sync with the click; Ensemble to fall apart due to click: the click track caused the ensemble to fall apart; Click sound itself is stressful: the click sound itself is stressful (too loud, prolonged exposure, painful to the ears); Requires intense concentration: requires intense concentration; Recording sessions: stress experienced during recording sessions; During practice: stress experienced during practice; Clicks hard to hear clearly: the click sound is hard to hear clearly; A sense of difficulty: a sense of difficulty or discomfort in keeping time with the click; My own playing is drowned out: my playing is drowned out by the click sound.

## Discussion

This study compared PRPPs between amateur and professional drummers in Japan, considering the distinct functional roles of each limb during performance. Specifically, we classified drummers into amateur and professional groups, examined their self-reported prevalence, affected body regions, and medical diagnoses of PRPPs, and identified the factors contributing to their development. We conducted a large-scale online survey of Japanese drummers.

The lifetime prevalence of self-reported PRPPs was 33.4% among amateur drummers, whereas it reached 66.1% among professional drummers. Regarding medical diagnoses, tenosynovitis was most frequently reported among amateurs, whereas MD was predominant among professionals. The distribution of affected body regions varied according to the performance level; amateurs most frequently reported problems in the upper limbs, particularly in the wrists, whereas professionals reported higher rates of problems in the right lower limb, followed by the left upper limb. Among drummers who reported PRPPs, 28.9% stated that they had received a medical diagnosis.

Risk factors associated with an increased probability of PRPPs included higher levels of performance activity, greater stress related to metronome click use, stronger tendencies toward perfectionism, and the experience of changing playing techniques, whereas greater stress resilience was associated with a decreased likelihood of PRPPs. A significant interaction was observed between music-activity level and stress resilience; in groups reporting higher levels, the protective effect of stress resilience was attenuated.

### Affected body region and diagnosis in Japanese drummers

A main findings of this study was that professional drummers most frequently reported PRPPs in the left upper and right lower limbs, which are primarily responsible for playing the snare and bass drums, and MD was the most commonly reported diagnosis in this group. Furthermore, the prevalence of MD among the professional drummers in this study was 8.95%. This finding contrasts that of Azar’s study of 831 drummers (80% professional drummers), which revealed no reported case of MD [13]. In a systematic review of multiple studies on MD, Rozanski et al. reported an incidence of 0.5–13% [34]. Among these studies focusing on musicians, particularly those in classical music, the most frequently reported prevalence rate was approximately 1%. Therefore, both the study by Azar and the present study may represent the extreme ends of the prevalence spectrum.

A possible explanation for the high prevalence rate observed in our study is the demanding technical requirements placed on Japanese professional drummers during stage performances and the associated psychological factors. Professional Japanese drummers are often required to perform with extreme temporal precision in synchronization with metronome clicks to match stage production, placing them under more stringent temporal constraints. Moreover, the right lower and left upper limbs, most frequently reported to be affected by professionals, form the rhythmic core of drumming performances, where the highest degree of temporal accuracy is required. Rozanski et al. identified fine motor demand as a key risk factor for MD, noting that the right hand is most frequently affected in keyboards and plucked string instruments (e.g., harps) [34]. Therefore, drummers may be exposed to excessive fine motor demands arising from extrinsic (synchronization with stage production) and intrinsic factors (precision in rhythmic patterns). Our logistic regression analysis further supported the role of psychological stress, identifying stress associated with the use of a metronome as a significant predictor of PRPPs.

Qualitative analysis of the comments of professional drummers regarding the click track revealed themes such as “Sense of isolation (being the only one hearing the click during performance)” and “Feeling caught in between the band’s natural groove and the click track.” Drummers are subjected to considerable stress because they are responsible for synchronizing with computers while simultaneously coordinating with other band members. Furthermore, the disappearance of the protective effect of stress resilience among nationally active drummers suggests a possible ceiling effect, whereby stress resilience is overwhelmed by the magnitude of stress. Taken together, these findings suggest that the increasing demand for synchronization with click tracks in modern stage performances may be a psychological trigger for MD among Japanese J-POP and J-Rock drummers.

The absence of MD reports in Azar’s study [13], in contrast to the high number of reports in our study, may be attributable to differences in the level of awareness of MD between North America and Japan. The technical requirements placed on North American professional drummers during stage performances may also be the same as those in Japan. What is the cause of this difference? One possibility is the difference in the recognition of MD. In Japan, a series of recent reports on MD among prominent drummers may have heightened the awareness of the condition among drummers. Conversely, Bledsoe et al., in a case series of American drummers with dystonia, reported that the average time from symptom onset to diagnosis spanned many years, attributing this delay to a lack of disease recognition among clinicians and delays in referral to specialists in neurology or movement disorders [18]. These differences in medical awareness and referral systems may have contributed to the discrepancy in reported cases. The second possible explanation involves cultural differences. Studies comparing East Asians, including Japanese, and Americans have shown that East Asians tend to evaluate themselves more negatively [35,36]. Such self-critical tendencies in East Asians may manifest as performance-related stress, potentially contributing to the increased incidence of MD in Japan.

In contrast, amateur drummers most frequently reported PRPPs on both wrists, with tenosynovitis as the most common diagnosis. In right-handed drummers, the hi-hat is typically played with the right upper limb, the snare drum with the left upper limb, and the bass drum with the right lower limb. Given the characteristics of playing the drum kit, the right upper limb used for the hi-hat was most frequently moved among right-handed performers. Higher levels of co-contraction in wrist flexors and extensors have been reported among amateur drummers [22]. Taken together, these findings suggest that the high incidence of wrist problems, with tenosynovitis being the most common diagnosis among amateurs, may result from a combination of less refined playing skills and high frequency of wrist movements.

Notably, our questionnaire did not determine whether participants used a right- or left-handed drum set or whether they played open-handed (left hand on the hi-hat, right hand on the snare) or closed-handed (right hand on the hi-hat, left hand on the snare). Therefore, it is important for future studies to discriminate between playing styles. Additionally, because the survey was disseminated with links to interview articles of SY and SF discussing PRPPs, drummers already experiencing PRPPs such as MD may have been more motivated to participate. Moreover, because the diagnoses were self-reported, there was no guarantee that they would be made by neurologists. Besides, the possibility of misdiagnosis at medical institutions must be considered.

### Lifetime prevalence of PRPPs as self-reported by the participants

In this study, 33.4% of amateur drummers and 66.1% of professional drummers self-reported PRPPs, with 28.9% of those affected having sought medical consultation. The prevalence of PRPPs among Japanese professional drummers was comparable to the 68% self-reported prevalence of PRMDs in Azar’s study [13] on predominantly North American drummers, 80% of whom were professionals, suggesting a similar rate of reporting. However, the rate of medical consultation was lower in our study (28.9%) than in Azar’s study (42%).

Similarly, a previous study on Japanese pianists reported that 44% of those with PRMDs sought medical consultation, whereas 57% of American piano students with PRMDs did so, indicating lower consultation rates among Japanese pianists [37]. Furuya et al. argued that several factors may underlie this low rate in Japan, including limited awareness of performing arts medicine among musicians and educators, a shortage of specialized clinicians and professional organizations in Asia, and cultural beliefs such as the notion that “pain should be endured” [37].

In addition, the study noted that self-treatment (e.g., icing or over-the-counter medication) is more common in Asia than in Western countries, which may further explain the lower consultation rates among Japanese pianists. Therefore, lower consultation rates among Japanese drummers compared to North American drummers may similarly be affected by the unique cultural background in Japan, as seen in pianists. Nevertheless, further investigation is warranted to clarify the relationship between cultural backgrounds and consultation rates among Japanese drummers.

### Risk factors: psychological and behavioral characteristics

A significant effect of perfectionism and change of technique on the risk of onset was confirmed. Perfectionism has been implicated as a risk factor for MD and PRMDs in numerous studies [3,27,38–40]. The results of the present study were consistent with those of previous studies. According to Hewitt and Flett, perfectionism can be conceptualized as comprising three dimensions: self-oriented, other-oriented, and socially prescribed perfectionism (SPP) [41].

Cruder et al. demonstrated that SPP, the externally imposed form of perfectionism typical of elite performers, constitutes a significant risk factor for PRMD onset among university music students [40]. In the present study, perfectionism was a significant predictor across all participants, including amateurs, without any interaction with the music-activity level. This suggests that forms of perfectionism other than SPP, such as self- or other-oriented perfectionism, may confer risks.

Our findings indicated that changes in playing techniques contributed to the onset of PRPPs. Previous studies have reported that technique modification increases susceptibility to PRPPs [42–44], and our results are consistent with these observations. Leijnse et al. hypothesized that when musicians acquire alternative motor patterns, practice beyond anatomical constraints can induce MD by overloading the related muscles. Although their study focused on pianists, string players, and wind players, similar mechanisms may be applicable in drummers. In cases where a change in playing technique contributes to the onset of PRPPs, the prognosis may improve if local inflammation is first controlled by medical interventions such as injections, followed by the collaboration of music educators and therapists to examine why excessive load is placed on the body, revise instrumental instruction, and implement physical training accordingly. However, it remains unclear whether the change in technique precipitated the onset or occurred as an adaptation to the early symptoms.

In the logistic regression model, variables such as total practice hours and lesson duration were not significant predictors. This finding is consistent with previous comparative studies of musicians with MD and healthy musicians, which also reported no significant differences in the total or daily practice time [9,45]. These observations suggest that temporal overuse alone does not lead to PRPP onset, emphasizing the need for multifactorial analyses that integrate the psychological, behavioral, and biomechanical risk dimensions.

### Clinical implication and application

A comparison of PRPPs among a wide range of individuals—from amateur to professional drummers—yielded important insights from clinical intervention and prevention perspectives. Based on a clinical intervention perspective, the identification of risk factors clarified background information that should be considered during diagnosis and treatment interventions (e.g., change of technique and perfectionism). These risk factors could serve as confounding variables in the evaluation of the therapeutic effects of treatments for PRMD, such as injections and medication prescriptions, and rehabilitation interventions. Furthermore, Détári [30] emphasized the importance of music educators participating in the development of new therapeutic approaches. Taken together, a comprehensive approach involving a multidisciplinary team—including orthopedic surgeons and neurologists who are direct specialists in PRPPs, therapists (e.g., physical therapists, occupational therapists), psychiatrists and psychologists to address perfectionist tendencies and depressive states, and music educators to consider performance perspectives—may lead to improved outcomes for musicians. Furthermore, a comprehensive approach incorporating these risk factors may enhance intervention effectiveness and contribute to relapse prevention.

Based on a prevention perspective, the reported incidence rates by body region indicate the sites where disorders are likely to occur due to long-term repetitive practice. The sites where PRPPs most frequently occur likely differ according to musical activity level. This study provides scientific evidence for revising practice and performance methods to prevent onset at each level; additionally, it provides evidence for proposing physical function training methods. Furthermore, the findings on lifetime prevalence and consultation rates are likely to highlight the issues regarding the present lack of medical and scientific support for drummers and issues in the social system.

### Limitations and future directions

This study has limitations that may have affected the interpretation of the results, as outlined below. Although the online survey format enabled the recruitment of participants from a wide geographical range, it also had certain limitations. A general limitation of online surveys is the potential concern regarding the reliability of responses and consistency of the data. In this study, a trap question was included to ensure data reliability. However, it was difficult to completely eliminate this issue. Additionally, the study data were collected through self-reporting, which may have been subject to cognitive biases or recall errors.

In future studies, it will be important to investigate cross-cultural differences in the impact of performance environments on PRPPs among drummers in other countries. Future research should further investigate the conditions of drummers and related factors in intercultural and international contexts by adopting a cross-cultural perspective.

## Data Availability

All data produced are available online at

https://osf.io/veck5/overview?view_only=76ec0f0948d14a5988b6b6c3b834f505

## Acknowledgments

We thank Dr. Nadia Azar for the discussions, Dr. Shiro Kumano and Dr. Aiko Murata for their help with the statistical analysis, and Natsuko Matsushita for depicting Figs 1, 2, and 4. We would like to thank Editage (www.editage.jp) for English language editing.

## Data availability

The datasets generated and/or analyzed during the current study and codes are available on the Open Science Framework ( https://osf.io/veck5/overview?view_only=76ec0f0948d14a5988b6b6c3b834f505).

### Supporting information

**S1 File. STROBE check list**

**S2 File. Questionnaire in Japanese (original).**

**S3 File. Questionnaire in English (translated).**

**S1 Fig. Original heatmap of frequency of affected body regions.**

## References

1. Guptill CA, Zaza C, Paul S. An occupational study of physical playing-related injuries in college music students. Med Probl Perform Art. 2000;15: 86–90. doi: 10.7939/R3K35MH83.

2. Lonsdale K, Boon OK. Playing-Related Health Problems Among Instrumental Music Students at a University in Malaysia. Med Probl Perform Art. 2016;31: 151–159. doi: 10.21091/mppa.2016.3028.

3. Elam T, Mowen S, Jonas C. Occupational injuries in musicians: A literature review. Mil Med. 2022;187: e619–e623. doi: 10.1093/milmed/usab499.

4. Zaza C, Charles C, Muszynski A. The meaning of playing-related musculoskeletal disorders to classical musicians. Soc Sci Med. 1998;47: 2013–2023. doi: 10.1016/s0277-9536(98)00307-4.

5. Altenmüller E. Focal dystonia: advances in brain imaging and understanding of fine motor control in musicians. Hand Clin. 2003;19: 523–38. doi: 10.1016/s0749-0712(03)00043-x.

6. Zaza C, Farewell VT. Musicians’ playing-related musculoskeletal disorders: an examination of risk factors. Am J Ind Med. 1997;32: 292–300. doi: 10.1002/(sici)1097-0274(199709)32:3<292::aid-ajim16>3.0.co;2-q.

7. Altenmüller E, Jabusch HC. Focal dystonia in musicians: phenomenology, pathophysiology, triggering factors, and treatment. Med Probl Perform Art. 2010;25: 3–9. doi: 10.21091/mppa.2010.1002.

8. Ackermann B, Driscoll T, Kenny DT. Musculoskeletal pain and injury in professional orchestral musicians in Australia. Med Probl Perform Art. 2012;27: 181–187. doi: 10.21091/mppa.2012.4034.

9. Moura RC, de Carvalho Aguiar PM, Bortz G, Ferraz HB. Clinical and Epidemiological Correlates of Task-Specific Dystonia in a Large Cohort of Brazilian Music Players. Front Neurol. 2017;8: 73. doi:10.3389/fneur.2017.00073.

10. Brandfonbrener AG. History of playing-related pain in 330 university freshman music students. Med Probl Perform Art. 2009;24: 30–36. doi: 10.21091/mppa.2009.1007.

11. Conti AM, Pullman S, Frucht SJ. The hand that has forgotten its cunning--lessons from musicians’ hand dystonia. Mov Disord. 2008;23: 1398–1406. doi: 10.1002/mds.21976.

12. Sandell C, Frykman M, Chesky K, Fjellman-Wiklund A. Playing-related musculoskeletal disorders and stress-related health problems among percussionists. Med Probl Perform Art. 2009;24: 175–180. doi: 10.21091/mppa.2009.4035.

13. Azar NR. Rates and Patterns of Playing-Related Musculoskeletal Disorders in Drummers. Med Probl Perform Art. 2020;35: 153–161. doi: 10.21091/mppa.2020.3020.

14. Azar NR. Injury Prevention Considerations for Drum Kit Performance. Front Psychol. 2022;13: 883279. doi: 10.3389/fpsyg.2022.883279.

15. Drummer M. Modern Drummer Education Team Weighs In On: Practicing With a Metronome. In: Modern Drummer Magazine [Internet]. 2012 May 8 [cited 31 Mar 2025]. Available: https://www.moderndrummer.com/2012/05/practicing-with-metronome/

16. Hawkins L. Investigating Musculoskeletal Pain Among Current Tertiary Drum Kit Players in Australia: A Mixed-Method Study Exploring Injury Risk Factors, Management and Prevention. Dissertation. Griffith University. 2015.

17. Lee A, Altenmüller E. Heavy metal curse: a task-specific dystonia in the proximal lower limb of a professional percussionist. Med Probl Perform Art. 2014;29: 174–176. doi: 10.21091/mppa.2014.3035.

18. Bledsoe IO, Reich SG, Frucht SJ, Goldman JG. Twelve Drummers Drumming… With Dystonia. Tremor Other Hyperkinet Mov. 2021;11: 6. doi:10.5334/tohm.577.

19. Margulis EH, Loui P, Loughridge D, editors. The Science-Music Borderlands: Reckoning with the past and imagining the future. The MIT Press; 2023. doi:10.7551/mitpress/14186.001.0001.

20. IFPI GLOBAL MUSIC REPORT 2025. [cited 2035 Mar 31]. Available: https://globalmusicreport.ifpi.org/

21. Ogawa Erika-shi ni Kiku, Nihon no Ongaku Shijō ni Tsuite Shitte Oku Beki Itsutsu no Koto [Five Things You Should Know About the Japanese Music Market: An Interview with Erika Ogawa]. [cited 2025 Mar 31]. Available: https://www.believe.com/newsroom/nippon-ongaku-shijo-nitsuite-tsu-koto (in Japanese).

22. Beveridge S, Herff SA, Buck B, Madden GB, Jabusch HC. Expertise-Related Differences in Wrist Muscle Co-contraction in Drummers. Front Psychol. 2020;11: 1360. doi: 10.3389/fpsyg.2020.01360.

23. Jabusch HC, Altenmüller E. Epidemiology, phenomenology, and therapy of musician’s cramp. Music, Motor Control and the Brain. unknown; 2006. pp. 265–282. doi: 10.1093/acprof:oso/9780199298723.003.0017.

24. Jacoby N, Polak R, Grahn JA, Cameron DJ, Lee KM, Godoy R, et al. Commonality and variation in mental representations of music revealed by a cross-cultural comparison of rhythm priors in 15 countries. Nat Hum Behav. 2024;8: 846–877. doi: 10.1038/s41562-023-01800-9.

25. Jacukowicz A. Psychosocial work aspects, stress and musculoskeletal pain among musicians. A systematic review in search of correlates and predictors of playing-related pain. Work. 2016;54: 657–668. doi: 10.3233/WOR-162323.

26. Foxman I, Burgel BJ. Musician Health and Safety: Preventing Playing-Related Musculoskeletal Disorders. AAOHN J. 2006;54: 309–316. doi: 10.1177/216507990605400703.

27. Détári A, Egermann H. Towards a holistic understanding of musician’s Focal Dystonia: Educational factors and mistake rumination contribute to the risk of developing the disorder. Front Psychol. 2022;13: 882966. doi: 10.3389/fpsyg.2022.882966.

28. Altenmüller E, Ioannou CI, Raab M, Lobinger B. Apollo’s curse: causes and cures of motor failures in musicians: a proposal for a new classification. Adv Exp Med Biol. 2014;826: 161–178. doi: 10.1007/978-1-4939-1338-1_11.

29. Ioannou CI, Furuya S, Altenmüller E. The impact of stress on motor performance in skilled musicians suffering from focal dystonia: Physiological and psychological characteristics. Neuropsychologia. 2016;85: 226–236. doi: 10.1016/j.neuropsychologia.2016.03.029.

30. Détári A. Treating the musician rather than the symptom: The holistic tools employed by current practices to attend to the non-motor problems of musicians with task-specific focal dystonia. Front Psychol. 2023;13:1038775. doi: 10.3389/fpsyg.2022.1038775.

31. Robin X, Turck N, Hainard A, Tiberti N, Lisacek F, Sanchez J-C, et al. pROC: an open-source package for R and S+ to analyze and compare ROC curves. BMC Bioinformatics. 2011;12: 77. doi: 10.1186/1471-2105-12-77.

32. Lüdecke D, Ben-Shachar M, Patil I, Waggoner P, Makowski D. Performance: An R package for assessment, comparison and testing of statistical models. J Open Source Softw. 2021;6: 3139. doi: 10.21105/joss.03139.

33. Braun V, Clarke V. Using thematic analysis in psychology. Qual Res Psychol. 2006;3: 77–101. doi: 10.1191/1478088706qp063oa.

34. Rozanski VE, Rehfuess E, Bötzel K, Nowak D. Task-specific dystonia in professional musicians. A systematic review of the importance of intensive playing as a risk factor. Dtsch Arztebl Int. 2015; 112: 871–877. doi:10.3238/arztebl.2015.0871.

35. Kitayama S, Markus HR, Matsumoto H, Norasakkunkit V. Individual and collective processes in the construction of the self: Self-enhancement in the United States and self-criticism in Japan. J Pers Soc Psychol. 1997;72: 1245–1267. doi: 10.1037/0022-3514.72.6.1245.

36. Peters HJ, Williams JM. Moving cultural background to the foreground: An investigation of self-talk, performance, and persistence following feedback. J Appl Sport Psychol. 2006;18: 240–253. doi: 10.1080/10413200600830315.

37. Furuya S, Nakahara H, Aoki T, Kinoshita H. Prevalence and causal factors of playing-related musculoskeletal disorders of the upper extremity and trunk among Japanese pianists and piano students. Med Probl Perform Art. 2006;21: 112–117. doi: 10.21091/mppa.2006.3023.

38. Bruyneel A-V, Stern F, Schmid A, Rieben N, James CE. Network analyses of physical and psychological factors of playing-related musculoskeletal disorders in student musicians: a cross-sectional study. BMC Musculoskelet Disord. 2024;25: 979. doi: 10.1186/s12891-024-08103-8.

39. Ioannou CI, Altenmüller E. Psychological characteristics in musician’s dystonia: a new diagnostic classification. Neuropsychologia. 2014;61: 80–88. doi: 10.1016/j.neuropsychologia.2014.05.014.

40. Cruder C, Soldini E, Gleeson N, Barbero M. Factors associated with increased risk of playing-related disorders among classical music students within the Risk of Music Students (RISMUS) longitudinal study. Sci Rep. 2023;13: 22939. doi: 10.1038/s41598-023-49965-7.

41. Hewitt PL, Flett GL. Perfectionism in the self and social contexts: Conceptualization, assessment, and association with psychopathology. J Pers Soc Psychol. 1991;60: 456–470. doi: 10.1037/0022-3514.60.3.456.

42. Revak JM. Incidence of upper extremity discomfort among piano students. Am J Occup Ther. 1989;43: 149–154. doi: 10.5014/ajot.43.3.149.

43. Sadnicka A, Kornysheva K, Rothwell JC, Edwards MJ. A unifying motor control framework for task-specific dystonia. Nat Rev Neurol. 2018;14: 116–124. doi: 10.1038/nrneurol.2017.146.

44. Leijnse JNAL, Hallett M, Sonneveld GJ. A multifactorial conceptual model of peripheral neuromusculoskeletal predisposing factors in task-specific focal hand dystonia in musicians: etiologic and therapeutic implications. Biol Cybern. 2015;109: 109–123. doi: 10.1007/s00422-014-0631-5.

45. Schmidt A, Jabusch HC, Altenmüller E, Kasten M, Klein C. Challenges of making music: what causes musician’s dystonia? JAMA Neurol. 2013;70: 1456–1459. doi: 10.1001/jamaneurol.2013.3931.

