## Supplementary material for "Playing-related physical problems: a large-scale online survey of professional and amateur Japanese drummers": S2 File Questionnaire in Japanese (original)_text-removed.docx

The text of the questionnaire (original, Japanese) has been removed due to the medRxiv’s policy that any language other than English should be removed.

If you want the original questionnaire in Japanese, please contact the corresponding author.
