## Supplementary material for "Playing-related physical problems: a large-scale online survey of professional and amateur Japanese drummers": S3 File Questionnaire in English (translated).docx

Study on Practice Methods of Drummers

and Physical Issues

The goal of this research is to clarify how drummers in Japan practice and what kinds of physical issues they deal with.

Please read the questionnaire instructions (Click Here). If you agree to participate, please select “Yes.”

All drummers are welcome to participate. Thank you in advance for your cooperation.

- Yes

If you do not agree, you may simply close your browser now.

Please indicate your sex.

- Male
- Female
- Non-binary/Third gender
- No response

Please indicate your age.

(Ex.) If you are 25 years old, use the cursor to select 25.

Age (years)

Which hand is your dominant hand?

- Right hand
- Left hand
- Both hands

What is your usual percentage of time spent on practicing with a matched grip and a traditional grip?

(Example: In the case of using the matched grip 70% of the time and the traditional grip 30% of the time, enter 70 in the “Matched grip” field and 30 in the “Traditional grip” field.)

- Matched grip
- Traditional grip
- Total

Select the genre of music that you play the most (multiple answers allowed).

If your answer is not provided as an option, select Other and type in the genre in the space provided. If you have more than one answer to type in, use a full-width comma “、” to separate your answers.

- Blues
- Classical
- Country
- Disco
- Hip-hop
- Jazz
- Metal
- Pop
- Reggae
- Rock
- Other (Type in your answer(s) in the following space)

Who is your favorite musician? If you have more than one answer to type in, use a full-width comma “、” to separate your answers.

(Example: aaaa、bbbb、cccc)

Which of the following applies to you?

- I am a perfectionist (I am not tolerant to even the slightest mistakes, tend to focus on flaws or past failures rather than successes, etc.)
- I tend to overfocus even on the small details (overfocusing)
- I am resilient to stress (stress resilience)
- I get enough sleep
- The genre of music I play is open to improvisation
- I often play to sound sources other than live performances (e.g. click tracks, soundtracks, programmed music, etc.)

1. Completely agree
2. Somewhat agree
3. Not sure
4. Somewhat disagree
5. Completely disagree

A problem with internet surveys is that some people lie, do not read the questions, or put no thought into their answers.

With this in mind, we would greatly appreciate if you could confirm that you have properly read these sentences.

If you have read them, do not respond to the questions below (in other words, do not click any of the options) and instead continue to the next page.

- I think so.
- I am inclined to think so.
- Neither
- I am not inclined to think so.
- I do not think so.

Please indicate the music-activity level that best applies to you, either pre-sent or past.

- Pro (For pay, national level)
- Pro (For pay, local level)
- Amateur (No pay, have performed in paid shows)
- Amateur (No pay, have performed in free shows）
- Amateur (No pay, enjoy playing on my own)
- No experience

How old were you when you first started playing drums?

(Ex.) If you began at 16 years of age, use the cursor to select 16.

・Beginning age (years)

How many years of experience do you have playing drums?

(Ex.) If the number of years is 15, use the cursor to select 15.

・Years of experience (number of years)

Have you played any other instruments?

- Yes
- No

Please enter the name of the instrument.

(Ex.) Piano

(Note) The same questions will be asked, so if you have experience with more than one instrument other than drums, please answer on a one-by-one basis.

Please indicate the age at which you began and how many years of experience you have.

(Ex.) If you began playing piano at 15 and have 3 years of experience, select 15 with the cursor for the age at which you started and 3 for years of experience.

- Beginning age (years)
- Years of experience (number of years)

Do you have experience playing another instrument?

- Yes
- No

Have you ever played a continuous sport?

- Yes
- No

Please indicate which sport.

(Ex.) Baseball

(Note) You will be asked the same questions, so if there is more than one sport, please answer on a one-by-one basis.

Please indicate the age at which you began and how many years of experience you have.

(Ex.) If you began playing baseball at 15 and have 3 years of experience, use the cursor to select 15 for the age at which you started and 3 for years of experience

- Beginning age (years)
- Years of experience (number of years)

Do you have experience with other sports?

- Yes
- No

Have you ever taken formal music lessons in a classroom setting or music school, a high school with a music program, or music college?

Please indicate the number of lesson hours per week taken in a music institution.

If the hours vary, please break them down by age.

(Ex.)

For example, imagine you took 3.5 hours per week at the age of 15 (the age at which you began) until 25 years of age, and from 26 to 40 years of age, you took 2 hours per week.

① “From age” in years will automatically be set.

・Age in years

・From age

・① Set automatically

② For “Until age,” use the cursor to select 25.

・Until age

・② Select 25

③ For lesson hours, use the cursor to select 3.5.

・Total number of lesson hours per week

・Hours

・③ Select 3.5 for 3.5 hours.

④ From the age of 26, the number of lesson hours changed, so select “Yes” below to add this change.

・Would you like to add a change to the lesson hours?

・Yes

・No

・④ Select “Yes” and proceed.

⑤ For the next “From age,” since 25 will be automatically set, use the cursor to select 26.

・Age

・From age

・⑤ Select 26.

⑥ For “Until age,” use the cursor to select 40.

・Until age

・⑥ Select 40.

⑦ Use the cursor to select 2 for the number of lesson hours.

・Total lesson hours per 1 week.

・Hours

・⑦ Select 2.

⑧ Continue to add changes until you reach the age lessons finished or the current age at which you are taking lessons.

(Note)

Here, only count practical lessons that use drums (such as ensemble practice and private drum lessons) and exclude hours of lessons that do not involve actual drumming (such as image training and music theory).

At what age did you start taking lessons?

・Age

・From age

・Until age

・Total number of lesson hours per 1 week

・Hours

Would you like to add any changes to these hours?

・Yes

・No

Please indicate the total number of hours per week you have practiced since beginning drumming until now. This does not include hours of practice obtained in lessons at a music education institution.

If the hours of practice vary, please break the hours down by respective age.

(Ex.)

For example, imagine that, from age 15 (the age you began drumming) to 25, you took 30 hours of lessons per week, and from 26 to 40 years of age, you took 15 hours per week.

①	The age in "From age” will be automatically set.

②	For “From age,” use the cursor to select 25.

③	For number of practice hours, use the cursor to select 30.

④	There is a change in the number of practice hours from the age of 26, so select “Yes” to add a change.

⑤	For the next “From age,” 25 will be automatically set, so use the cursor to select 26.

⑥	For “Until age,” select 40.

⑦	For hours practiced, select 15.

⑧	Continue to add changes until you reach the age you stopped practicing or the current age at which you are practicing.

(Note)

From here, count only actual hours of practice involving drums, including with a band or on your own. This does not include practice hours that do not involve actual drumming even if they include exercises that involve moving the hands and feet.

- Age
- From age
- Until age
- Total number of hours per week you have practiced
- Hours

Would you like to add any changes to these hours?

Please indicate other ways you often practice outside of lessons.

(Ex.) Band practice, rudiment practice, etc.

Have you ever made a significant change to your play style?

・Yes

・No

Around what age did this change occur?

(Ex.) Around 17 years old, I changed from being self-taught to using the Moeller method.

In each situation below, how often have you used click tracks?

If a situation is not listed, please enter it into the “Other” field.

(Ex.) If used 50% of the time when practicing, use the cursor to select 50%.

・Practice (%)

・Recording (%)

・Live performance (%)

・Other (%)

What is the maximum and minimum tempo you usually play at?

(Sound source)

Ref.: 60 BPM

Ref.: 120 BPM

Ref.: 180 BPM

・Maximum BPM

・Minimum BPM

What type of click track do you use? (Multiple answers allowed)

If your answer is not provided as an option, select Other and type in the type of click track in the space provided.

・Cowbell

・Wood sound

・Electronic sound

・Rim shot

・Human voice

・Other

How do you listen to these click tracks (multiple choices possible)?

If a situation is not listed below, enter it into the “Other” field.

・During practice

・During recording

・During live performance

・Other

・In-ear headphones

・Headphones

・Directly from a speaker or metronome

・Do not use

Please indicate the volume at which you listen to click tracks per situation.

Referring to the figure below, please give your subjective judgment of the volume of the click tracks when playing.

If a situation is not listed below, enter it into the “Other” field.

(Ex.) If you listen at 60 dB (decibels) when practicing, use the cursor to select 60 in the “Practice” field.

・Practice (%)

・Recording (%)

・Live performance (%)

・Other (%)

Have you ever felt stress by using a click track?

If you have, describe the situation where you felt stressed in the space below.

(Example: I felt stressed when only I had to listen to a click track during a live performance. It required a lot of concentration, because being out of sync with the click track means that I am not in sync with the lighting, video, and other production elements.)

・Yes

・No

Have you felt disorientation, discomfort, or bodily pain that occurs regularly when performing?

・Yes

・No

When and in what situation did you experience this, and what were the symptom(s) involved?

(Example: For XX years (or XX months) since age XX, I felt stiffness when I tried XX (strokes, etc.). I experience uncontrolled movement. I feel pain.

I started feeling dizziness from being nervous during performance since around age XX. I get strange cold sweats.)

(Note: Since a similar question is asked after this, if you have multiple symptoms that you are concerned about, prioritize and provide those that are causing problems with playing drums.)

Using the chart below, please select the parts where symptoms appear (multiple choices possible).

If the location of, for example, dizziness or extreme outbreaks of sweat, are difficult to pinpoint, please continue on without selecting anything.

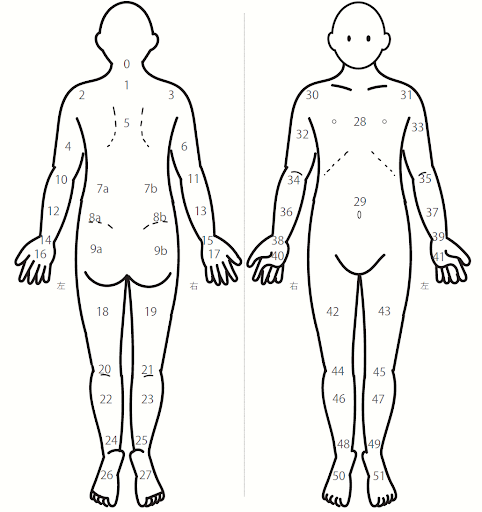

Have you consulted a medical professional about your symptoms and been given a diagnosis?

・Yes

・No

What diagnosis were you given?

(e.g., Myofascial pain syndrome, Tenosynovitis, Focal dystonia, etc.)

At what age did you receive the diagnosis?

・Age (years)

Are there any other parts where you have experienced issues?

・Yes

・No

In the future, we plan to investigate in detail the body’s movements and use of muscles when playing drums. If you would like to participate, please provide your email address.

If you wish to participate in this survey only and have your response record deleted at a later date, please provide a nickname of your choice that is three letters or longer so that you can be identified.

If you close out of the browser without entering your email address and nick-name, the questionnaire response results will all be lost and will not be recorded.

(Note)

①	Please refrain from choosing strings of letters or numbers such as “AAAAA” or “11111”.

②	Please make note of your email or nickname as they will be needed should you wish to have your response record deleted.

③	If you provide your email address, we may contact you about future experiments. Your email may also be used for data identification in the case that you wish to have your response record deleted. Your email address will not be used for any reasons other than those listed above.

This concludes the questionnaire.

If you agree to having your answers recorded, please select “Yes” and continue.

・Yes

If you do not agree, please close your browser without continuing. Your data will not be recorded.

Thank you very much for your help on the questionnaire.

Your responses have been recorded.
