## Supplementary figures and images for "Playing-related physical problems: a large-scale online survey of professional and amateur Japanese drummers"

### S1 Fig Original heatmap.png

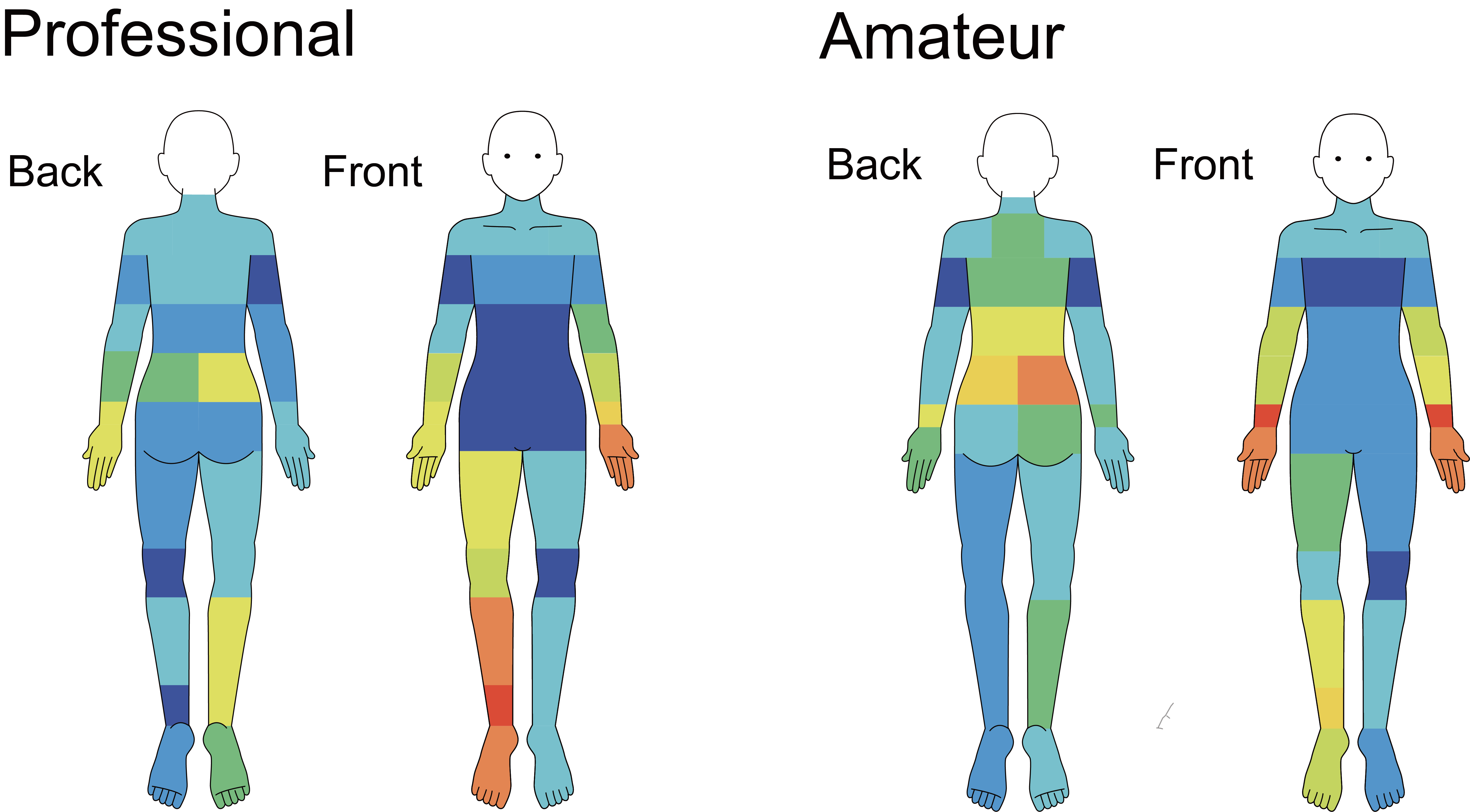
